# Inequity in the face of success: Understanding geographic and wealth-based equity in success of facility-based delivery for under-5 mortality reduction in six countries

**DOI:** 10.1101/2023.05.29.23290665

**Authors:** Jovial Thomas Ntawukuriryayo, Amelia VanderZanden, Alemayehu Amberbir, Alula Teklu, Fauzia Akhter Huda, Mahesh Maskey, Mohamadou Sall, Patricia J Garcia, Raj Kumar Subedi, Sayinzoga Felix, Lisa R Hirschhorn, Agnes Binagwaho

**Author notes:** Corresponding author and email: Jovial Thomas Ntawukuriryayo. **Author contact emails:** Amelia VanderZanden, Alemayehu Amberbir, Raj Kumar Subedi, Mahesh Maskey, Patricia J Garcia, Mohamadou Sall, Felix Sayinzoga, Alula M. Teklu, Fauzia Akhter Huda, Lisa R Hirschhorn, Agnes Binagwaho. **Trial Registration: N/A**.

## Abstract

**Background:** Between 2000-2015, many low- and middle-income countries (LMICs) implemented evidence-based interventions (EBIs) known to reduce under-5 mortality (U5M). Even among LMICs successful in reducing U5M, this drop was unequal subnationally, with varying success in EBI implementation. Building on mixed methods multi-case studies of six LMICs (Bangladesh, Ethiopia, Nepal, Peru, Rwanda, and Senegal) leading in U5M reduction, we describe geographic and wealth-based equity in facility-based delivery (FBD), a critical EBI to reduce neonatal mortality which requires a trusted and functional health system, and compar<u>e</u> the implementation strategies and contextual factors which influenced success or challenges within and across the countries.

**Methods:** To obtain equity gaps in FBD coverage and changes in absolute geographic and wealth-based equity between 2000-2015, we calculated the difference between the highest and lowest FBD coverage across subnational regions and in the FBD coverage between the richest and poorest wealth quintiles. We extracted and compared contextual factors and implementation strategies associated with reduced or remaining inequities from the country case studies.

**Results:** The absolute geographic and wealth-based equity gaps decreased in three countries, with greatest drops in Rwanda – decreasing from 50% to 5% across subnational regions and from 43% to 13% across wealth quintiles. The largest increases were seen in Bangladesh – from 10% to 32% across geography – and in Ethiopia – from 22% to 58% across wealth quintiles. Facilitators to reducing equity gaps across the six countries included leadership commitment and culture of data use; in some countries, community or maternal and child health insurance was also an important factor (Rwanda and Peru). Barriers across all the countries included geography, while country-specific barriers included low female empowerment subnationally (Bangladesh) and cultural beliefs (Ethiopia). Successful strategies included building on community health worker (CHW) programs, with country-specific adaptation of pre-existing CHW programs (Rwanda, Ethiopia, and Senegal) and cultural adaptation of delivery protocols (Peru). Reducing delivery costs was successful in Senegal, and partially successful in Nepal and Ethiopia.

**Conclusion:** Variable success in reducing inequity in FBD coverage among countries successful in reducing U5M underscores the importance of measuring not just coverage but also equity. Learning from FBD interventions shows the need to prioritize equity in access and uptake of EBIs for the poor and in remote areas by adapting the strategies to local context.

## Introduction

For over five decades, reducing under-5 mortality (U5M) has been one of the greatest public health challenges globally (1) with the investment in the Millennium Development Goals (MDGs) accelerating needed declines in U5M. Some of the success has been through the development and implementation of evidence-based interventions (EBIs) delivered by health systems. These EBIs have included vaccinations, effective facility- and community-based care for common illnesses for children under 5 (U5), and work to improve maternal and neonatal care (Supplement Table 1) (2–7). Despite significant advances in decreasing U5M globally, work remains for many countries to ensure that progress in lowering U5M, including neonatal mortality, is equitable across geography and wealth (8,9).

There have been many countries which were more successful in decreasing U5M between 2000-2015 compared with their regional and economic peers supported through implementation of these EBIs. We focused on six of these countries, Ethiopia, Peru, Bangladesh, Nepal, Rwanda, and Senegal, for multi-country mixed methods implementation research case studies to understand the implementation strategies and contextual factors in achieving or falling short in national coverage of these EBIs (2–7). These countries had relative reductions of U5M which varied from 57% in Nepal to 74% in Rwanda. Similar reductions in neonatal mortality (NMR) took place, though at lower relative percent, ranging from 41% in Senegal to 72% in Bangladesh (Supplement Table 2) (10–21). Building from these case studies, we describe geographic and wealth-based equity in the coverage of facility-based delivery (FBD) and compare the implementation strategies and contextual factors which influenced success or challenges within and across the countries. We chose this EBI as it is a critical intervention to reduce NMR, one which requires a complexity of skills and a trusted and functional health system. This work is important to inform policymakers and implementers of strategies needed to identify and address barriers and leverage facilitators to not only achieve coverage but ensure that this progress is equitable across their countries and populations.

## Methods

### Facility-Based Delivery Coverage

For each of the selected countries, we extracted data from the available Demographic and Health Survey (DHS) reports from 2000 (baseline) and 2015 (endline) on FBD coverage for the national level, the subnational regions with the highest and the lowest coverage, and for the poorest and richest quintiles.

### Case studies

The details of the methodology and specific country case studies are described elsewhere in detail (4,22–27). Briefly, these case studies were conducted using an implementation research framework we developed to understand how the six countries implemented health system-delivered EBIs known to reduce amenable U5M (27). We used explanatory mixed methods to gather data from DHS, key informant interviews, and a desk review, to understand implementation of EBIs including FBD, identify strategies the countries used for EBI implementation, and local contextual factors which influenced implementation outcomes including reach, equity, and acceptability (27). The key informants were identified in collaboration with in-country partners to capture experience and insights from a range of stakeholders including Ministry of Health (MOH) decision-makers and policymakers, implementers, and donors and implementing partners active during the study period. Key informant interviews (KIIs) were analyzed using thematic content approach to extract contextual factors, implementation strategies, and implementation outcomes for each of the countries, supplemented by the desk review results.

### Equity analysis

For this study, we calculated the absolute equity gaps (absolute value difference) and change between 2000-2015 in FBD coverage based on WHO recommendations (28), as follows:

1. Economic equity: difference in the FBD coverage between the richest and poorest wealth quintiles.
2. Geographic equity: difference between the highest and lowest FBD coverage across subnational regions (28).

We identified strategies which were implemented to address potential challenges to ensure equitable coverage of FBD, including addressing the existing contextual factor(s) across wealth quintiles and subnational regions (27). Based on evidence from the case studies, implementation strategies were categorized as associated with improved equity, partially successful, not successful, or no evidence of association with improved equity. Contextual factors identified in the case studies were also categorized as barriers, facilitators, both, or not found in influencing success in equitable coverage.

### Ethical considerations

The Rwanda National Ethics Committee and Northwestern University determined the research to be non-human subjects study. Each country case study was reviewed and approved by in-country Ethics Review Committees (Ethiopia, PM23/281; Peru, 104276; Bangladesh, PR-18074; Nepal, 165-2018; Senegal, SEN18/33; Rwanda, 132/RNEC/2017). Interview participants provided verbal informed consent after receiving clear information about the goals and structure of the research.

## Results

### Coverage of facility-based delivery

We found that all six countries had increased FBD coverage at the national level in the DHS survey closest to 2015 as compared to baseline (Table 1; Figure 1). At baseline, FBD coverage was below 50% for all countries except Peru and Senegal, where coverage was close to 60%. In the endline survey, there was an absolute increase ranging from 12-65 percentage points and the mean coverage was above 80% for Peru and Rwanda, though it remained below 50% for Ethiopia and Bangladesh.

**Figure 1.**
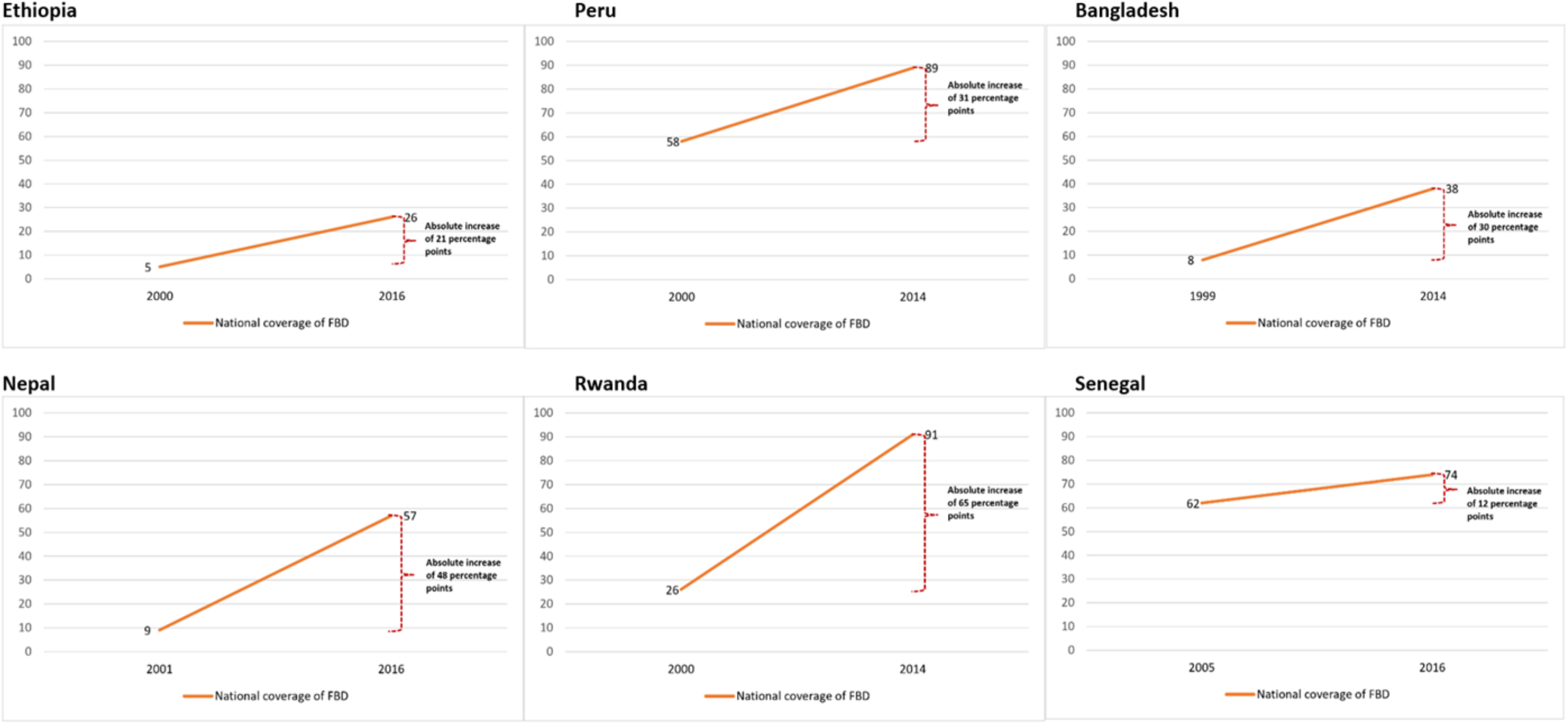
National change in facility-based delivery coverage between 2000 and 2015 (10–14,16–21,47)

**Table 1.**
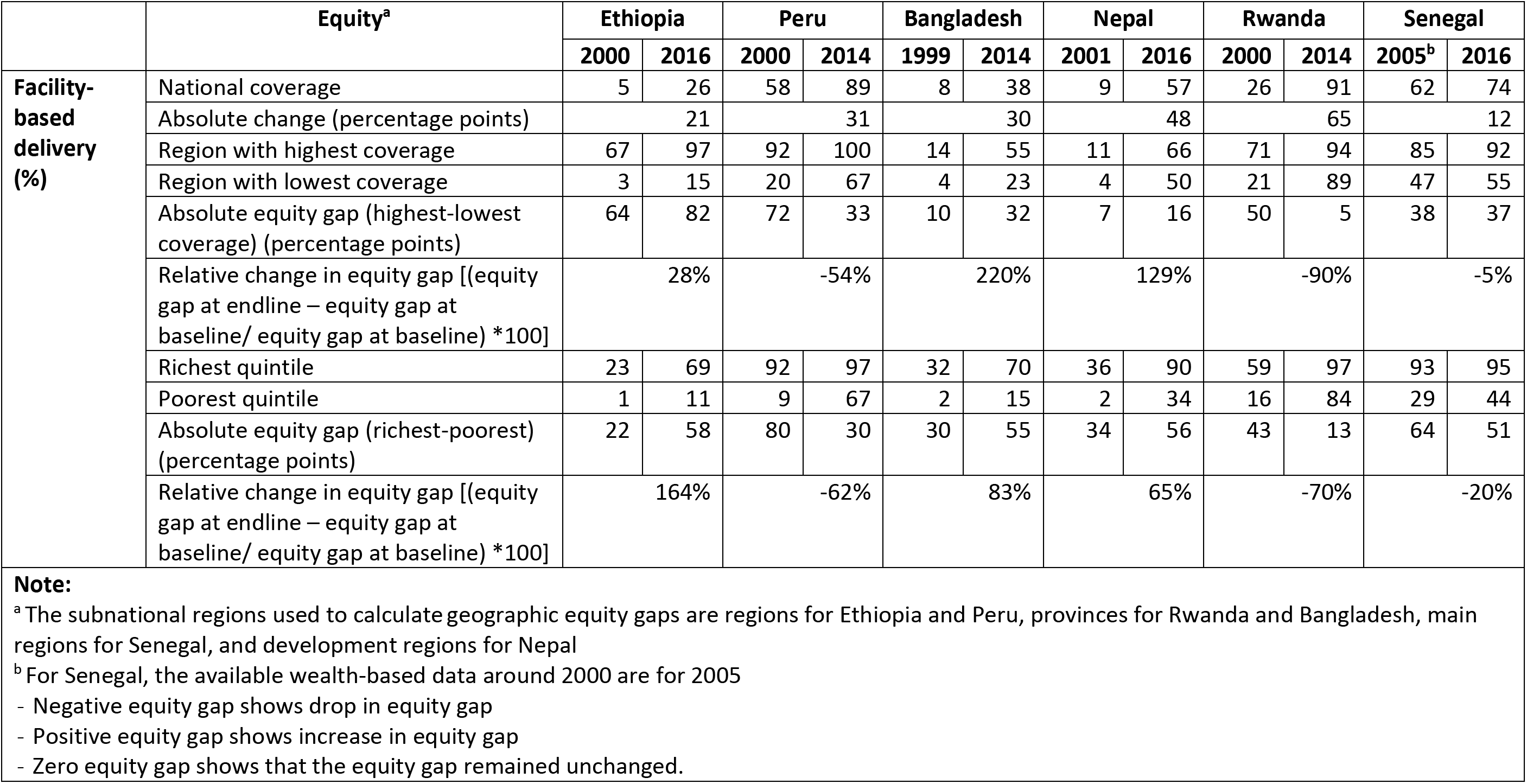
Coverage and change in equity gaps in facility-based delivery across wealth quintiles and subnational regions (10–14,16–21,28,47)

|  | Equity<sup>a</sup> | Ethiopia |  | Peru |  | Bangladesh |  | Nepal |  | Rwanda |  | Senegal |  |
| --- | --- | --- | --- | --- | --- | --- | --- | --- | --- | --- | --- | --- | --- |
|  |  | 2000 | 2016 | 2000 | 2014 | 1999 | 2014 | 2001 | 2016 | 2000 | 2014 | 2005<sup>b</sup> | 2016 |
| Facility-based delivery (%) | National coverage | 5 | 26 | 58 | 89 | 8 | 38 | 9 | 57 | 26 | 91 | 62 | 74 |
|  | Absolute change (percentage points) | 21 |  | 31 |  | 30 |  | 48 |  | 65 |  | 12 |  |
|  | Region with highest coverage | 67 | 97 | 92 | 100 | 14 | 55 | 11 | 66 | 71 | 94 | 85 | 92 |
|  | Region with lowest coverage | 3 | 15 | 20 | 67 | 4 | 23 | 4 | 50 | 21 | 89 | 47 | 55 |
|  | Absolute equity gap (highest-lowest coverage) (percentage points) | 64 | 82 | 72 | 33 | 10 | 32 | 7 | 16 | 50 | 5 | 38 | 37 |
|  | Relative change in equity gap [(equity gap at endline – equity gap at baseline/ equity gap at baseline) *100] | 28% |  | -54% |  | 220% |  | 129% |  | -90% |  | -5% |  |
|  | Richest quintile | 23 | 69 | 92 | 97 | 32 | 70 | 36 | 90 | 59 | 97 | 93 | 95 |
|  | Poorest quintile | 1 | 11 | 9 | 67 | 2 | 15 | 2 | 34 | 16 | 84 | 29 | 44 |
|  | Absolute equity gap (richest-poorest) (percentage points) | 22 | 58 | 80 | 30 | 30 | 55 | 34 | 56 | 43 | 13 | 64 | 51 |
|  | Relative change in equity gap [(equity gap at endline – equity gap at baseline/ equity gap at baseline) *100] | 164% |  | -62% |  | 83% |  | 65% |  | -70% |  | -20% |  |
| **Note:**<br><sup>a</sup> The subnational regions used to calculate geographic equity gaps are regions for Ethiopia and Peru, provinces for Rwanda and Bangladesh, main regions for Senegal, and development regions for Nepal<br><sup>b</sup> For Senegal, the available wealth-based data around 2000 are for 2005<br>- Negative equity gap shows drop in equity gap<br>- Positive equity gap shows increase in equity gap<br>- Zero equity gap shows that the equity gap remained unchanged. |  |  |  |  |  |  |  |  |  |  |  |  |  |

### Change in equity in facility-based delivery

Coverage increased in all subnational regions. However, differences in rates and change varied within the six countries. At baseline, the absolute equity gap based on geography ranged from 7 percentage points in Nepal to 72 percentage points in Peru, while at the endline, the gap ranged from 5 percentage points in Rwanda to 82 percentage points in Ethiopia. Between the baseline and endline, this absolute equity gap decreased for Rwanda (from 50% to 5%) and Peru (from 72% to 33%), remained almost unchanged for Senegal, and increased somewhat in Ethiopia, Nepal, and Bangladesh (Table 1).

Coverage also increased across all wealth quintiles, but it remained lower in the poorest quintiles compared to the richest quintiles regardless of country. The baseline absolute wealth gap ranged from 22 percentage points in Ethiopia to 80 percentage points in Peru. At the endline, this gap ranged from 13 percentage points in Rwanda to 58 percentage points in Ethiopia. The wealth equity gap decreased in Peru, Senegal, and Rwanda, ranging from an absolute decline of 13 percentage points in Senegal (from 64% to 51%) to 50 percentage points in Peru (from 80% to 30%). The wealth equity gap widened in other countries, ranging from the absolute increase of 22 percentage points (from 34% to 56%) for Nepal to 36 percentage points (from 22% to 58%) for Ethiopia (Table 1).

### Contextual factors

In all the countries, there were contextual factors that either facilitated or hindered the reduction of inequities in FBD coverage across the wealth quintiles and/or subnational regions (Table 2). Facilitators across all countries included leadership commitment, culture of data use, existing community health workers (CHWs), and focus on female empowerment.

**Table 2.** List of contextual factors across the six countries.

| +: Facilitated equity in FBD coverage -: Barrier to equity in FBD coverage +/-: Both facilitator and barrier<br>N/I: Not identified as influencing factor |  |  |  |  |  |  |  |
| --- | --- | --- | --- | --- | --- | --- | --- |
| Contextual Factor | Countries |  |  |  |  |  |  |
|  | Senegal | Bangladesh | Nepal | Rwanda | Ethiopia | Peru | Decision |
| Conflict | - | - | N/I | N/I | - | N/I | - |
| Cultural beliefs | N/I | - | N/I | N/I | - | N/I | - |
| Culture of data use | + | + | + | + | + | + | + |
| Economic growth | + | + | + | + | + | + | + |
| Existing community health system | + | + | + | + | + | + | + |
| Female empowerment status | + | +/- | + | + | +/- | + | +/- |
| Financial commitment to the health sector | N/I | N/I | N/I | + | N/I | N/I | + |
| Geography | - | - | - | +/- | - | - | +/- |
| Health insurance | +/- | +/- | - | + | + | + | +/- |
| Health systems strength and strengthening | +/- | N/I | +/- | N/I | +/- | + | +/- |
| Language barrier | N/I | - | N/I | N/I | N/I | N/I | - |
| Leadership commitment | + | + | + | + | + | + | + |
| Poverty | - | - | - | - | - | - | - |

The existing national leadership facilitated implementation of EBIs targeted at improving maternal and child health. In Peru, for example, maternal and child health was among key priorities in the national Roundtable for the Fight Against Poverty (26). It allowed delivery of health services including FBD across different settings by prioritizing the poorest populations. All six countries also had a culture of data use with a well-structured system of data management and reporting at national level. The culture of data use at the national level helped in the understanding of improvements needed to address inequity of coverage of maternal and child health EBIs including FBD across different subnational areas and wealth quintiles. An example of the culture of data use is the Government of Senegal’s recognition of low FBD coverage in the lowest wealth quintile and rural areas, which resulted in the introduction of a free delivery policy to facilitate access to FBD for the poorest population. Another example is the Safe Motherhood multimedia campaign implemented in response to low FBD coverage data in rural areas and some regions in Peru. The program targeted to improve women’s awareness about benefits of FBD and encouraged them to deliver in health facilities. In addition, CHWs were in charge of basic childcare and health promotion activities including FBD, helping to reach more vulnerable populations across different geographic settings including remote areas. For female empowerment, improvements in education access and decision-making power benefited many women from different geographic areas and socio-economic classes in all the countries. This was associated with increased awareness about maternal and child health care including benefits of FBD. However, some gaps were described by the KIIs. For example, female empowerment remained lower in rural settings than urban areas, particularly in hard-to-reach areas. Finally, the economic development seen in all countries was associated with increased access to and availability of FBD. For instance, a Peru Health Reform Program (locally known as *Programa de Apoyo a la Reforma del Sector Salud*, or PARSALUD) financed from the Public Treasury, but mainly from the Inter-American Development Bank and the World Bank, to improve maternal and child health including FBD, primarily in the poorest regions.

Facilitating factors which were specific to only some countries included health insurance systems which ensured that the most vulnerable population would have access to health services (Rwanda and Peru) as well as performance contracts of local leaders (Rwanda). In Rwanda, around 90% of the general population used community-based health insurance (CBHI), introduced in 2000. The government covered the annual subscription for the poorest along with their 10% of cost for reimbursement at each point of care. Pregnant women with CBHI cards from poor families could deliver in health facilities without out-of-pocket cost; pregnant women with some financial means paid only 10% of the cost. This removed the financial burden to health services for households. In Peru, free health insurance for children between the ages of 3-17 years old who were enrolled into public schools was introduced in 1997. In 1998, the country also established a maternal and child health insurance program which covered costs of health care including FBD for women from low-income families. This coverage contributed to increased use of health facilities for delivery for women in the poorest quintile. In 2001, the two insurance programs were combined into a public Integral Health Insurance (known as *Seguro Integral de Salud* [SIS]) to improve financial access to health care services for the mothers and children under 18 within the poor population.

Prominent barriers across most countries included geographic obstacles. Some pregnant women living in remote areas had difficulties with reaching health facilities for delivery due to long travel distance and lack of transport. In Ethiopia, for example, only 5% of those who lived at a distance greater than five kilometers away from a health center delivered in health facilities. Although CHWs delivered care from nearer facilities called health posts, they did not provide FBD services. Thus, even if the health posts improved access for other services, FBD access and utilization remained low for those living farther away from facilities which provided FBD. In Bangladesh, the limited road infrastructure in rural areas made seeking care at health facilities for poor and pregnant women living in rural settings very difficult. Mountainous and hard-to-reach areas of Sylhet had the lowest FBD coverage among other divisions, an example of this challenge. While these countries all experienced economic development, which facilitated FBD expansion and access, persistent poverty in sub-populations was also a shared barrier.

We also found some country-specific barriers which influenced subnational and/or wealth-based equity. This included regional language barriers (Bangladesh), cultural beliefs (Ethiopia, Bangladesh), subnational areas with low female empowerment (Bangladesh), and areas of civil unrest (Senegal, Bangladesh, Ethiopia). Bangladesh had a language barrier at the subnational level as most health professionals in Sylhet, a rural division, came from other regions to address health care worker shortages and did not speak the local language of the Sylheti. Cultural beliefs affected FBD in rural regions of Ethiopia, where some women considered it unnecessary, inconvenient because they were not allowed to be accompanied by family members in health facilities as they were in their homes, and it was not in their customs. KIs explained that pastoralist regions have sparsely distributed populations with strong cultural beliefs, and the lack of cultural birthing adaptation contributed significantly to the subnational inequity.

Subnational female empowerment was challenging in Bangladesh, and specifically in Sylhet, where it was linked to cultural and religious beliefs. This contextual factor influenced women’s decision-making power, including the ability to go out of the family to seek services such as health care including FBD. Political instability within regions in some countries also limited access to health facilities for patients. For example, the Casamance region in southern Senegal experienced conflict between 1992-2014, with subsequent challenges of easy access to health facilities for delivery in the region. Similar challenges also occurred in the Chattogram division in Bangladesh due to ongoing civil unrest since the late 1980s, as well as in the Afar and Tigray regions in Ethiopia at the Ethiopian-Eritrean border since 1998.

### Implementation strategies

We found that the six countries implemented a range of strategies to address contextual factors that influenced access and equity in FBD coverage across geography and wealth (Table 3). These strategies commonly focused on the health system including financial and physical access to, and acceptability of, FBD, although how they were implemented varied. The strategies focused on improving acceptability included leveraging existing CHW programs (all), adaptation of existing clinical protocols to reflect cultural practices (Peru), and integrating CHWs focusing solely on maternal health into existing community health programs (Rwanda and Senegal). For example, in Peru many women from rural areas preferred to deliver in vertical position. In response to the identified barrier to acceptability of the standard protocols, Peru adapted clinical protocols and delivery rooms to the cultural preferences to increase willingness of these women to deliver in health facilities, contributing to the improvement of subnational equity in FBD (26).

**Table 3.**
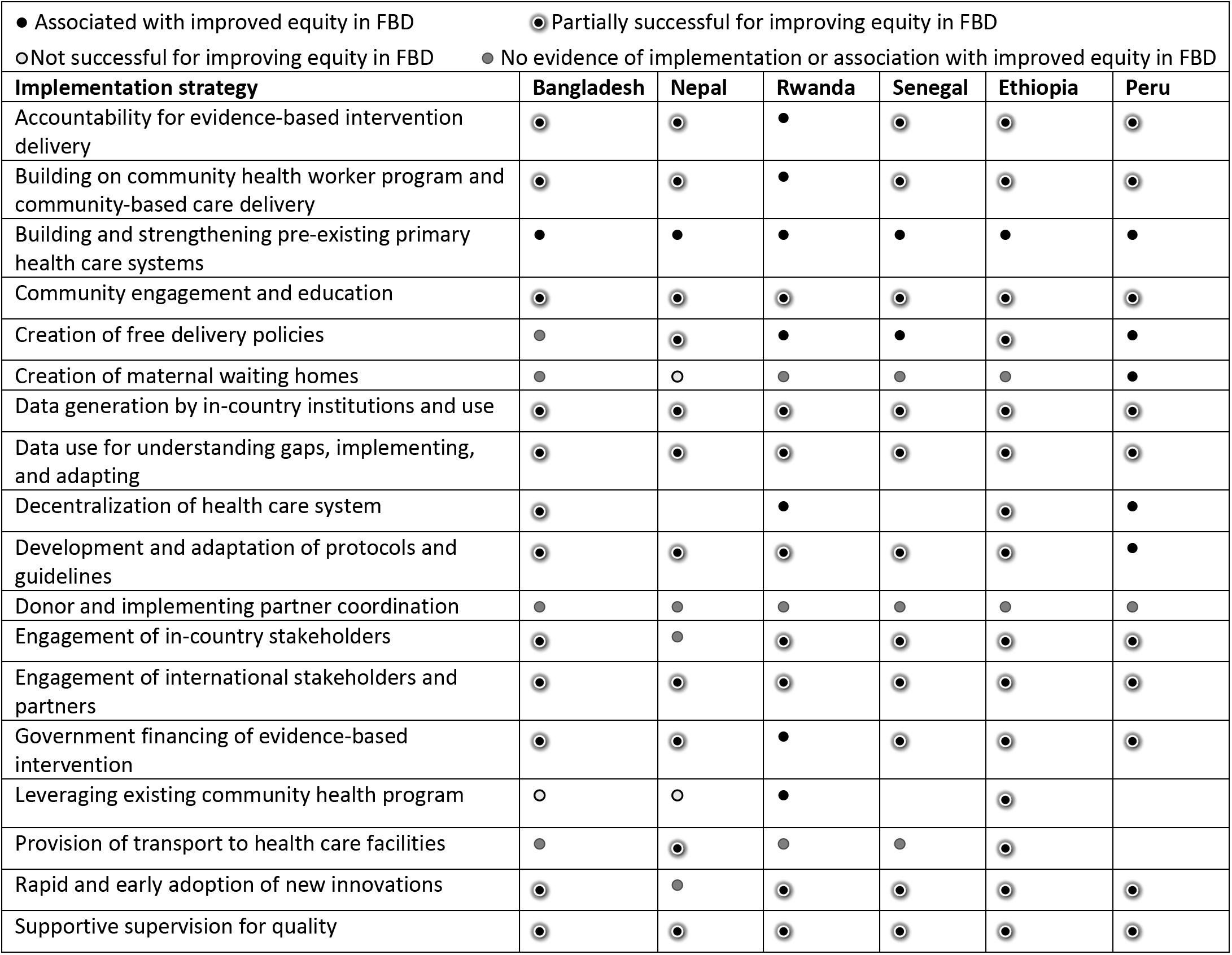
Implementation strategies for reducing inequity in facility-based delivery.

The six countries leveraged existing CHW programs differently. For instance, Rwanda added a CHW in charge of maternal health to the existing CHW program in each village. This individual raised awareness about the benefits of FBD, accompanied pregnant women to health facilities for delivery when necessary, and called for medical emergency support for pregnant women in danger – particularly those living in remote areas. In Senegal, some women – particularly those in rural and remote areas – had customs of hiding early pregnancies. This resulted in delays in seeking health care and medical advice, including FBD. In response, the country introduced a new cadre of CHWs, the *bajenou gokh,* who were typically respected older women from the community. The *bajenou gokh* helped with detecting when women were in early pregnancy and encouraged them to seek care, including FBD, in a timely way. Health extension workers in Ethiopia identified early pregnancies during home visits, provided antenatal care, facilitated patient referral to nearby health facilities, and supervised the Women’s Development Army. The Women’s Development Army was a cadre in charge of educating pregnant women, particularly those from generally poorer population groups in rural areas, about the benefits of health facility-delivered care services including FBD, and accompanying these women to health facilities for delivery. The decline of home deliveries was among indicators considered in the performance contracts of the health extension workers, especially those in pastoralist communities.

Strategies which were part of broader health system strengthening and also helped improve physical access to health facilities for delivery included decentralization of health facilities and deployment of health workers, for example, in more remote areas (Rwanda, Nepal, and Peru); establishment of maternal waiting homes near health facilities for pregnant women from distant locations who had difficulties with reaching the facilities while in labor (Peru, Ethiopia, and Nepal); and distribution of ambulances in rural areas (Ethiopia). To improve geographic access to health facilities for people from remote areas including pregnant women, Rwanda decentralized the health care system, building new health centers with a target of at least one per administrative sector serving a population of around 25,000 people. Rwanda also began to establish one health post per administrative cell serving approximately 5,000 people in cells without existing health centers. This strategy, along with the allocation of trained health professionals by prioritizing rural areas, contributed to improved access to FBD services across all areas. Ethiopia similarly decentralized health facilities, with each health center serving 25,000 people and each health post providing care services to 5,000 people. In Peru, as pregnant women in remote areas had challenges in accessing FBD once in labor, the country established maternal waiting homes close to health facilities where women nearing their due date could stay along with their preferred relatives. These waiting homes were not preferred at the beginning, but later used after being adapted to the women’s preferences (26). Similarly, Nepal created maternal waiting homes for pregnant women from more remote settings to stay near health facilities, where they would receive urgent medical support, while waiting for their due date. However, these waiting homes were not used because local women did not want to spend days in those maternity homes as they had other responsibilities in their homes. Ethiopia leveraged the maternity waiting homes which existed since the 1970s to accommodate pregnant women from remote settings to deliver in health facilities. However, the utilization of the homes was low because of negative perception, for instance, insufficient space and latrines and the community perceived them as the only option for the poorest.

To reduce the distance to health facilities and improve access to FBD for pregnant women, Bangladesh created community clinics staffed with community health care providers and community-based skilled birth attendants. These providers delivered maternal and child health services including FBD in remote and hard-to-reach areas. However, weak monitoring and supervision as well as the high workload of the community-based skilled birth attendants were major challenges.

Financial access to health facilities for delivery was improved through the provision of free or subsidized delivery services for poor pregnant women (Senegal, Ethiopia, and Nepal) and through provision of transport incentives to pregnant women from low-income families living in the poorest districts (Nepal). Senegal improved access to health facilities for delivery through the creation of a free delivery and Cesarean policy, which allowed pregnant women from the poorest regions and hard-to-reach areas to receive free normal delivery services at health posts and health centers as well as delivery by Cesarean section at regional hospitals. Nepal also implemented a free delivery policy and provided financial incentives to facilitate transport to health facilities for pregnant women from poor families living in hard-to-reach areas. However, the women tended to bypass peripheral health facilities including health centers and health posts in preference for delivery in hospitals, possibly due to low readiness of Basic Emergency Obstetric and Newborn Services at the peripheral facilities. In response to limited means of transport to health facilities for delivery for pregnant women living in remote areas, Ethiopia implemented a free normal delivery policy and distributed ambulances across all its regions to enable transport of women to health facilities for delivery for free. However, the follow up on implementation of these strategies was weak at regional levels.

## Discussion

Although national findings on reducing U5M showed that the six countries outperformed their peers in LMICs (2–7), FBD coverage still remained below 50% for some countries, with variations across wealth quintiles and subnational regions (2–7). None of the six countries we studied were able to eliminate the equity gap in FBD coverage, though Rwanda had the greatest reduction of the gap across the wealth quintiles and subnational regions.

In addition to lower national coverage at the endline remaining below 50%, Bangladesh and Ethiopia were the only countries to see increases in the absolute equity gap across the subnational regions and wealth quintiles, with a small number of areas contributing to these gaps. While FBD coverage increased from 9% to 57% in Nepal, that country also was not able to decrease the equity gap. These results are similar to the study of FBD by Victora et al (2017), which reported absolute equity gaps tending to be the highest when national FBD coverage is around 50% (29). In addition, when national coverage is 60% or above, the equity gaps tend to be the lowest, although the most vulnerable group continued to lag behind (29).

We found leadership commitment and accountability and culture of data use to be important contextual factors. These factors were reflected in the inclusion of maternal and child health services including FBD into the national Roundtable for the Fight Against Poverty in Peru (7) and maternal and child health indicators including FBD into performance contracts of local leaders in Rwanda (2,30). These approaches encouraged national and local leaders to follow up on the progress of improving FBD coverage at subnational levels. Butrick et al. (2014) also found the leadership commitment effective, for example when Nigeria’s president demonstrated political will through the 2012 announcement of funds to support social safety net programs which included maternal and child health indicators, focusing on areas with the greatest need. The study further highlighted the Presidential Initiative for Safe Motherhood launched by the president of Malawi which focused on banning traditional birth attendance, expanding maternal waiting homes, and introducing community midwives to ensure safe delivery for pregnant women from resource-limited settings (31). Improvement in female empowerment in the six Exemplar countries was also a strong contextual factor facilitating increased FBD coverage, as awareness about FBD benefits and financial status increased among women. These results are similar to findings by Asseffa, Bukola, and Ayodele (2016), where FBD was associated with higher female education levels and financial means to afford FBD in a preferred health facility (32).

As Rwanda and Peru either created or redesigned health insurance systems that considered removing the financial burden of health services including FBD on the poorest population, the FBD gap across wealth quintiles reduced in these countries (33,34). This benefit was also found by a study conducted in Tanzania, where pregnant women from poor families were given free maternal and child health insurance to allow them to receive health services, including safe delivery in health facilities, until three months after delivery. This strategy greatly contributed to increases in FBD coverage among the poorest population (35).

The wide equity gaps that existed at baseline were mainly related to limited access to health facilities due to lack of financial means to pay for FBD or transport to health facilities. Geographic barriers restricted pregnant women in remote settings from accessing health facilities in the six countries, and these barriers worsened if the women were poor. Other studies have similarly found that long distance to health facilities and lack of transport means were associated with reduced FBD for women from poor population groups, particularly those from rural remote settings (36,37).

The findings of language and cultural context as a barrier were also noted in Guatemala by Ishida and colleagues (2012), who found lower access to quality care services including delivery services when local women spoke a language different from the one spoken by health care providers (38). Cultural beliefs on the importance of home-based delivery as well as the long distance to health facilities in some areas of Ethiopia may be associated with increased equity across geography and wealth quintile. Similar cultural beliefs and long distance to health facilities were also reported as restricting Maasai populations in Kenya from use of health facilities for safe delivery (39). The conflict in the Casamance region in the south of Senegal may explain the unchanged subnational equity gap in FBD due to limited access to health facilities to seek care including delivery. A study conducted for the populations of Uganda and Egypt found that coverage of FBD tended to be lower in areas with civil unrest as access to transport and delivery services became limited (40,41).

The adaptation of delivery protocols to the culturally-sensitive practices in Peru may also explain the reduced equity gap across subnational regions. Adapting this delivery protocol is in line with a systematic review which found that delivery standards need to be adapted to local culture to be more acceptable and successful (42). The CHWs specific to maternal health who provided health education and follow up on each pregnancy (Senegal and Rwanda) may explain the reduced equity gap across subnational regions. Research in India found that in a quite similar program, where CHWs were given mobile phones containing maternal and child health messages to regularly educate pregnant women and particularly those in remote settings about benefits of seeking health care, this strategy was associated with increased FBD (43).

The creation of maternal waiting homes in Peru may be associated with the reduced equity gap in FBD across both wealth quintiles and subnational regions. One of the reasons the waiting homes were successful in Peru but not the other countries we examined could be for the extra effort to adapt the homes to the women’s preferences (26). Similar waiting homes were welcomed by communities in Malawi as they were constructed in different sites near health care facilities in 2012 to support pregnant women from poor families living in areas distant from delivery facilities (31).

Slight reductions in the wealth-based equity gap in Senegal may be explained by the free delivery policies, as evidenced from a study conducted in Burkina Faso which found that subsidization or removal of delivery costs resulted in reduced absolute value differences in the coverage between the richest and poorest quintiles (44). Although a study conducted in Kenya found that output-based voucher programs to motivate the most vulnerable women to seek health services including FBD could result in increases in FBD among the poorest (45), the free delivery policies implemented along with transport facilitations in Nepal and Ethiopia were not able to reduce the equity gaps, possibly due to weak follow-up at subnational levels.

Our study had a number of limitations. We focused only on FBD; strategies and contextual factors for other EBIs may differ. However, across the case studies, a number of these limitations were common regardless of setting or intervention. They included inability to establish causality or attribution between a number factors and strategies to equity change; limited data on outcomes such as acceptability across countries even though coverage can reflect access, acceptability, or both; limited number of key informants; and inability to target women who had FBD or home delivery to ask about their experience. Another limitation was that we did not account for population sizes across subnational regions or wealth quintiles while measuring inequity, with use of regression-based inequality measures such as slope index of inequality and the relative index of inequality. We were also not able to measure the quality of FBD and home delivery subnationally including safety of delivery interventions, timely response to pregnancy-related problems, and people-centeredness, and so can only reflect on coverage, not effective coverage (46). Future studies would take these limitations into consideration.

## Conclusion

While the six Exemplar countries saw an overall decrease in U5M and NMR at national levels, equity gaps in FBD coverage remained across wealth quintiles and subnational regions throughout the study period. Some countries made substantial reductions in equity gaps in FBD coverage, while others were still struggling with narrowing this gap by the end of 2015. Learning from FBD as an EBI also shows the need to increase efforts in prioritizing equity in access and uptake of EBIs for the poor and subnational regions in rural and remote areas by adapting the strategies to local context. The experience from these countries in the contextual factors and implementation strategies offers potentially transferable knowledge for policymakers and decision-makers within these countries and other countries working to decrease the inequities in FBD across wealth quintiles and subnational regions.

## Data Availability

The datasets used and/or analyzed during the current study are available from the corresponding author on reasonable request.

## List of abbreviations

CBHI: Community-based health insurance
CHW: Community health worker
DHS: Demographic and Health Survey
EBI: Evidence-based intervention
FBD: Facility-based delivery
KI: Key informant
KII: Key informant interview
LMIC: Low- and middle-income country
MDGs: Millennium Development Goals
MOH: Ministry of Health
NMR: Neonatal mortality
PARSALUD: Programa de Apoyo a la Reforma del Sector Salud
SIS: Seguro Integral de Salud
U5: Under 5
U5M: Under-5 mortality
WHO: World Health Organization

## Declarations

### Ethics approval and consent to participate

The Rwanda National Ethics Committee and Northwestern University determined the research to be non-human subjects study. Each country case study was reviewed by and received approval from in-country Ethics Review Committees (Ethiopia, PM23/281; Peru, 104276; Bangladesh, PR-18074; Nepal, 165-2018; Senegal, SEN18/33; Rwanda, 132/RNEC/2017). Interview participants provided verbal informed consent after receiving clear information about the goals and structure of the research.

### Consent for publication

Not applicable

### Author’s contributions

JTN, LRH, and AB made substantial contributions to the design of this work.

JTN, AB, and LRH contributed to the data analysis.

JTN, LRH, AV, AA, and AB interpreted the results to form the manuscript.

JTN, LRH, AV, AT, FAH, MM, MS, PLG, RKS, SF, AA, and AB contributed to writing and revision of the manuscript have approved the final version.

## Acknowledgments

We want to acknowledge and thank the key informants and other stakeholders who provided essential information, historical perspectives and narratives, and feedback on our findings, ensuring we captured the most accurate reflection possible on these countries’ journeys to reducing under-5 mortality.

## Author’s information

Corresponding author: Jovial Thomas Ntawukuriryayo

## Supplement tables

**Supplement Table 1.** List of neonatal evidence-based interventions.

| Period of risk | Evidence-based intervention |  |
| --- | --- | --- |
| **Preconception** | Folic acid supplementation |  |
| **Antenatal** | Tetanus vaccination |  |
|  | Malaria prevention and treatment | Intermittent presumptive treatment<br>Insecticide-treated bed nets |
|  | Iodine supplementation (in endemic iodine deficient settings) |  |
|  | 4 or more antenatal visits (ANC4) |  |
|  | Prevention and treatment of preeclampsia and eclampsia | Antihypertensive treatment for severe hypertension<br>Magnesium sulfate<br>Early delivery |
| **Intrapartum** | Antibiotics for preterm premature rupture of membranes |  |
|  | Corticosteroids for preterm labor |  |
|  | C-section for breech or obstructed labor |  |
|  | Active management of delivery (including partograph) |  |
|  | Clean delivery practices (incl. clean cord-cutting) |  |
|  | Trained birth attendant |  |
|  | Facility-based delivery |  |
|  | Basic emergency obstetric and newborn care (BEmONC) |  |
|  | Comprehensive emergency obstetric and newborn care (CEmONC) |  |
|  | Timely transport for higher level care for mother |  |
| **Postnatal** | Newborn resuscitation |  |
|  | Immediate breastfeeding |  |
|  | Prevention and management of hypothermia | Immediate drying and wrapping<br>Delayed bathing<br>Skin-to-skin<br>Baby warming |
|  | Kangaroo care for LBW/prematurity |  |
|  | Timely transport for higher level care for mother |  |
|  | Post-partum visits to identify danger signs and provide active referral |  |
|  | Antibiotics for suspected or confirmed infection |  |
|  | Surfactant therapy for respiratory distress syndrome and prematurity |  |
|  | Neonatal intensive care units (equipped, trained staff, standards and protocols established and followed) |  |

**Supplement Table 2.** Change in under-5 mortality and neonatal mortality in six countries (10–21)

| Country | Ethiopia |  | Rwanda |  | Senegal |  | Peru |  | Bangladesh |  | Nepal |  |
| --- | --- | --- | --- | --- | --- | --- | --- | --- | --- | --- | --- | --- |
|  | 2000 | 2016 | 2000 | 2015 | 1999 | 2015 | 2000 | 2014 | 2000 | 2014 | 2001 | 2016 |
| Under-5 mortality<br>(deaths per 1000 live<br>births) | 166 | 67 | 196 | 50 | 145 | 59 | 60 | 23 | 110 | 30 | 91 | 39 |
| Relative change (%) | **-60** |  | **-74** |  | **-59** |  | **-62** |  | **-73** |  | **-57** |  |
| Neonatal mortality<br>(deaths per 1000 live<br>births) | 58 | 29 | 44 | 19 | 39 | 23 | 23 | 12 | 50 | 14 | 39 | 21 |
| Relative change (%) | **-50** |  | **-57** |  | **-41** |  | **-48** |  | **-72** |  | **-46** |  |

